# Safety and Outcomes of Valproic Acid in Subarachnoid Hemorrhage Patients: A Retrospective Study

**DOI:** 10.1101/2024.09.09.24313246

**Authors:** Matthew Cobler-Lichter, Kushak Suchdev, Hayley Tatro, Ava Cascone, Joanna Yang, Janice Weinberg, Mohamad K Abdalkader, Hormuzdiyar H Dasenbrock, Charlene J Ong, Anna Cervantes-Arslanian, David Greer, Thanh N Nguyen, Ali Daneshmand, David Y Chung

## Abstract

**Background and Purpose:** Animal studies have suggested that valproic acid (VPA) is neuroprotective in aneurysmal subarachnoid hemorrhage (SAH). Potential mechanisms include an effect on cortical spreading depolarizations (CSD), apoptosis, blood-brain barrier integrity, and inflammatory pathways. However, the effect of VPA on SAH outcomes in humans has not been investigated.

**Methods:** We conducted a retrospective analysis of 123 patients with nontraumatic SAH. Eighty-seven patients had an aneurysmal source and 36 patients did not have a culprit lesion identified. We used stepwise logistic regression to determine the association between VPA and the following: delayed cerebral ischemia (DCI), radiographic vasospasm, and discharge modified Rankin Scale (mRS) score > 3.

**Results:** All 18 patients who received VPA underwent coil embolization of their aneurysm. VPA use did not have a significant association with DCI on adjusted analysis (Odds Ratio, OR = 1.07, 95% CI: 0.20 – 5.80). The association between VPA use and vasospasm was OR = 0.64 (0.19 – 1.98) and discharge mRS > 3 was OR = 0.45 (0.10 – 1.64). Increased age (OR = 1.04, 1.01 - 1.07) and Hunt and Hess (HH) grade > 3 (OR = 14.5, 4.31 – 48.6) were associated with an increased likelihood for poor discharge outcome (mRS > 3). Younger age (OR = 0.96, 0.93 - 0.99), mFS score = 4 (OR = 4.14, 1.81 – 9.45), and HH grade > 3 (OR = 2.92, 1.11 – 7.69) were all associated with subsequent development of radiographic vasospasm. There were no complications associated with VPA administration.

**Conclusion:** We did not observe an association between VPA and the rate of DCI. There may have been a protective association on discharge outcome and radiographic vasospasm that did not reach statistical significance. We found that VPA use was safe and is plausible to be used in a population of SAH patients who have undergone endovascular treatment of their aneurysm. Larger, prospective studies are needed to determine the effect of VPA on outcome after SAH.

## Introduction

Aneurysmal subarachnoid hemorrhage (SAH) has an estimated annual incidence of 12 out of 100,000 people in the United States.^1,2^ SAH has an in-hospital mortality rate of 20%^3^ and a 30-day rate of 35% and is a major cause of long term disability.^1^ Poor outcomes following SAH are caused not only by the initial hemorrhage, but also by the sequelae known as cerebral vasospasm and delayed cerebral ischemia (DCI).^4–6^

The pathophysiology of DCI has yet to be fully understood. It was long believed that vasospasm was the direct cause for DCI. Recent studies have shown that the cause of DCI is likely multifactorial and includes cerebral vascular dysfunction, microthrombosis, cortical spreading depolarizations (CSD) and neuroinflammation.^7^

CSDs have recently emerged as a potential contributor to secondary brain injury following SAH and clinical studies have shown strong associations between CSDs and poor outcome in this population.^8^ Therefore, drugs used to block CSD are an appealing therapeutic approach to prevent and treat DCI. Multiple animal and clinical studies have investigated prevention of CSD-mediated secondary injury following SAH. These studies have included a) vasodilating drugs used to transform spreading ischemia to hyperemia^9^, b) IV fluids and nimodipine to alter microcirculation to prevent spreading ischemia^10^, c) use of the phosphodiesterase-3 inhibitor cilostazol to shorten the duration of spreading ischemia^11^ and d) valproic acid (VPA) to reduce injury from CSDs.^12^

VPA is a known inhibitor of CSDs^12,13^ and improves neurological outcome in rodent models of SAH.^12,13^ However, to our knowledge, no human studies have been performed to determine the relationship between VPA and outcome in patients following SAH. VPA is sometimes given to hospitalized patients to manage headache, agitation, or seizures. Therefore, we evaluated the effect of VPA given for these other indications on functional outcome in patients with SAH, as well as the potential effect of VPA in reducing vasospasm and DCI.

## Methods

The reporting of this study adheres to the guidelines contained in the STROBE statement.^14^ All methods were carried out in accordance with relevant guidelines and regulations. This study was approved by the Institutional Review Board at Boston Medical Center including a waiver of informed consent (BMC IRB H-40681). We retrospectively reviewed prospectively collected records of 169 consecutive patients with subarachnoid hemorrhage (age ≥ 21 years) from the Get with the Guidelines stroke registry at Boston Medical Center between 2014 and 2020. Patients with non-traumatic SAH were included in the study. Exclusion criteria included traumatic, convexal, or sulcal SAH, intracerebral hemorrhage, brain death or transition to comfort measures only (CMO) within 24 hours of admission, leaving AMA during the hospitalization, and the presence or history of cerebral arteriovenous malformation (AVM), reversible cerebral vasoconstriction syndrome (RCVS), or other vasculopathies (Figure 1). Each individual patient’s chart was reviewed by a neurointensivist (AD, KS, or DYC) to ensure that only appropriate patients were included, and to adjudicate data abstraction.

**Figure 1.**
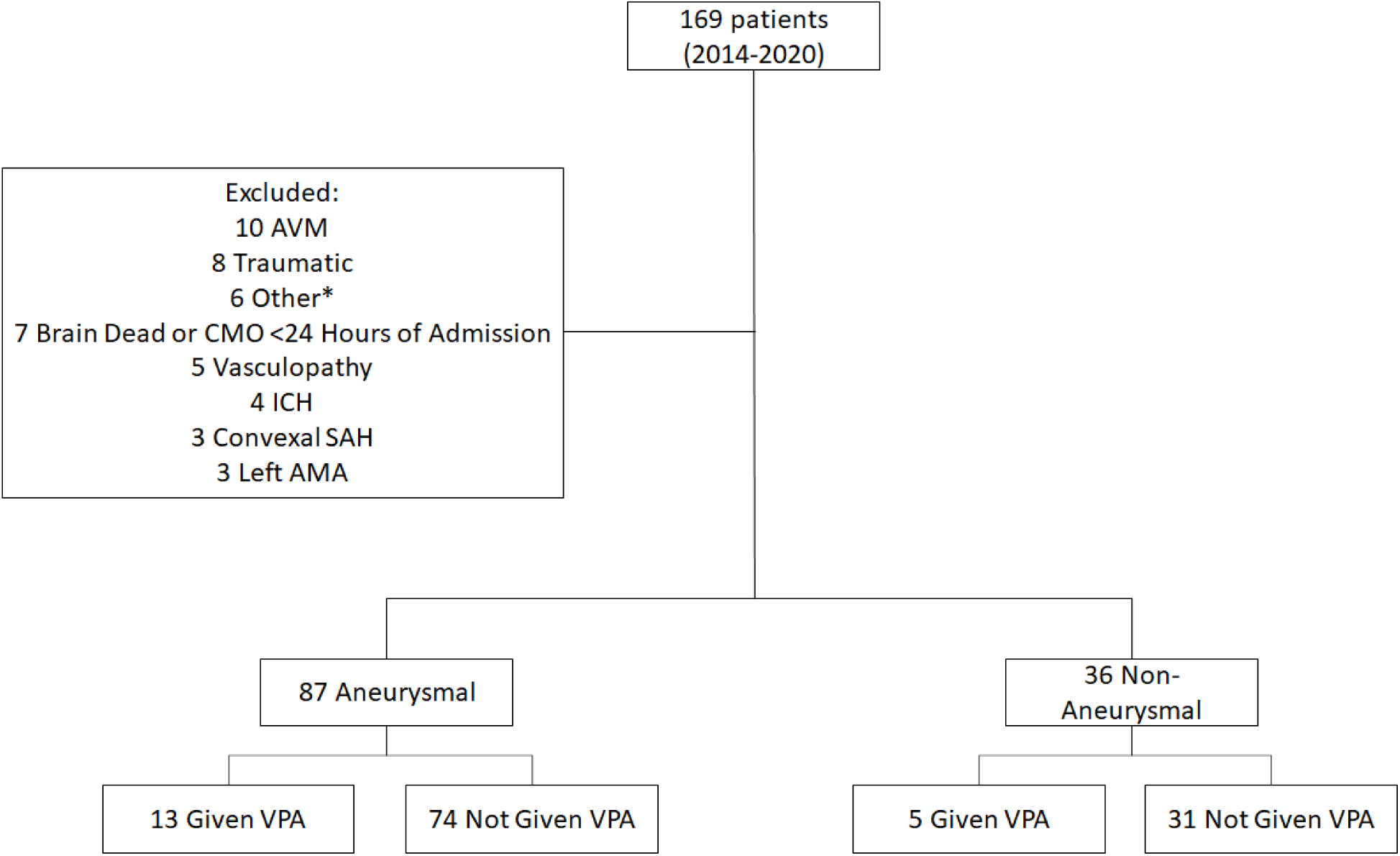
Distribution of patients that were considered for study inclusion. AVM = arteriovenous malformation, CMO = comfort measures only, RCVS = reversible cerebral vasoconstriction syndrome, VPA = valproic acid. *One intradural hematoma, one admitted for re-rupture of aneurysm that initially ruptured a month prior, one already included as previous record, one subdural hemorrhage, one cavernoma, and one sulcal SAH.

All cases of radiographic vasospasm and DCI were adjudicated among three neurointensivists (AD, KS, and DC). Radiographic vasospasm was defined by the presence of one or both of the following: moderate to severe vasospasm on CT or digital subtraction angiography or TCD mean flow velocities > 120 cm/sec in the middle cerebral artery and Lindegaard ratios > 3. DCI was defined by the presence of one or more of the following: new focal neurologic deficit lasting at least 1 hour, Glasgow coma scale (GCS) decline in the total of significant magnitude (≥2 points), or radiologic signs of infarction. Two subcategories of DCI were defined and included in the analysis, which include clinical deterioration (CD) attributed to DCI and cerebral infarction (CI) attributed to DCI.^4,5,15^

### Data Collection and Outcomes

VPA was given to hospitalized patients with SAH to manage headache, agitation, or seizures. We recorded patient demographics, admission Hunt and Hess (HH) grade, World Federation of Neurological Surgeons (WFNS) Grade, aneurysm treatment modality (e.g. coiling vs. clipping), aneurysm location and size, presence of EVD, VPA data including duration and dose, and the admission modified Fisher Scale (mFS) score. Race and ethnicity are reported in accordance with the AMA Manual of Style.^16^ The main outcomes of interest were rates of radiographic vasospasm, DCI, and discharge outcome mRS > 3 vs. <4.

### Safety Review of VPA

We conducted a review of all patients who received VPA to determine how many patients experienced VPA-related complications. VPA has been associated with thrombocytopenia, reduction of platelet aggregation, transaminitis, hyperammonemia (normal below 35 μmol)^17^, and elevated amylase and lipase.^18,19^ We reviewed clinical notes and relevant lab values including platelet counts and liver function tests (LFTs) before, during, and after VPA administration. Thrombocytopenia was defined as mild (100-149 × 10^^9^/L platelet count), moderate (50-99 × 10^9^/L), or severe (< 50 × 10^9^/L).^20^

We also required a 50% reduction from baseline platelet count after VPA was administered to diagnose VPA-associated thrombocytopenia. We excluded other etiologies such as heparin-induced thrombocytopenia, severe sepsis, and disseminated intravascular coagulopathy. The list of complications searched for included hyperammonemia, thrombocytopenia, bleeding disorders, pancreatitis, and liver function abnormalities.

### Statistical Methods

Unadjusted analyses were conducted using Chi Square tests for categorical data and t-tests for continuous normally-distributed data (Table 2). A Wilcoxon rank sum test was used for continuous non-parametric data. A value of p < 0.05 was used as the predefined threshold for statistical significance. Stepwise logistic regression analyses were conducted to determine the association between VPA use and DCI, vasospasm, and mRS > 3. We selected final logistic regression models by testing all predictor variables against each of the three outcome variables and selecting predictor variables with p-values that were below 0.10. The use of VPA was included in the models despite failure to meet the p-value cutoff. The final logistic regression model for DCI, vasospasm, and discharge mRS > 3 included age, HH grade > 3, mFS score = 4^21^.^22^, and VPA use. The only predictor variable that met the p-value cutoff for DCI was mFS score = 4, likely due to the fact that there were only 12 participants who experienced DCI. Despite this, the predictor variables were kept the same in each model with each given outcome variable for consistency. Statistical analyses were performed using JMP Pro 15 and SAS.

## Results

The final analyzed study population was 123 patients, of whom 87 had aneurysmal SAH and 36 did not have a culprit lesion (Figure 1). Fifteen percent of patients (18/123) were given VPA. Baseline demographics were similar between the VPA and non-VPA groups except no surgically clipped patients received VPA (Table 1). The median total dose of VPA over the course of hospitalization was 55.8 mg/kg (range: 5.9 - 831.8) and the median time to start VPA was post-bleed day 3 (PBD, range: 1-13). The median daily dose of VPA was 10.4 mg/kg/day (range: 3-41.5). The median duration of VPA administration was 5.5 days (range: 1 – 20). Figure 2 includes additional details about day of initiation, dosing, duration, and VPA levels.

**Figure 2.**
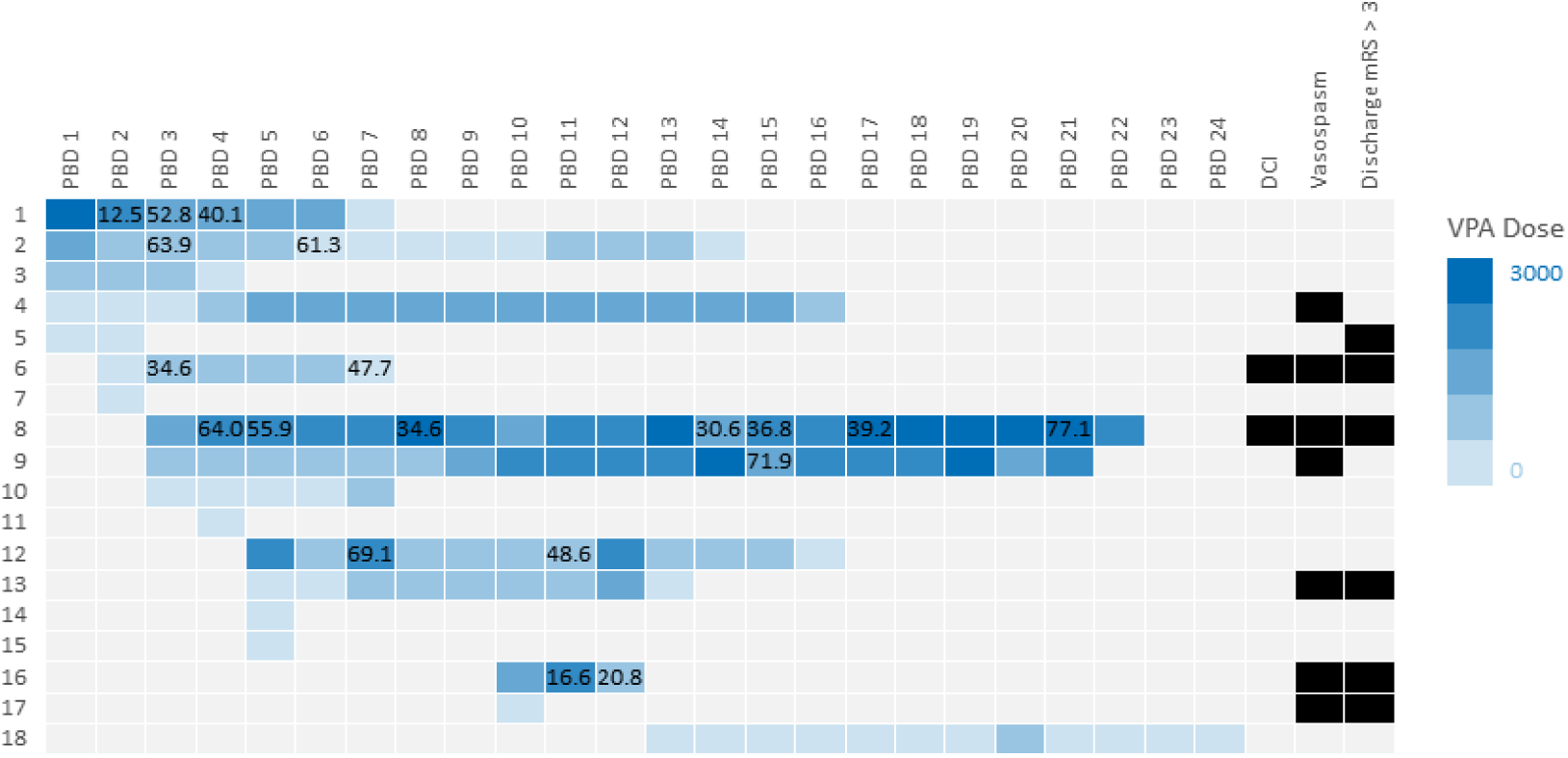
Dose and Duration of VPA administration with outcomes for each patient who received VPA. Blood levels of VPA are displayed in each box for which the data was available (mcg/mL).

**Table 1.** Baseline and Clinical Characteristics of patients with SAH in the VPA and non-VPA groups.

| Characteristic | No VPA Administered | VPA Administered | P Value |
| --- | --- | --- | --- |
| Number | 105 | 18 |  |
| Mean Age (yrs) | 57.2 ± 14.6 | 53.1 ± 15.6 | 0.30 |
| Female (%) | 71 (67.6) | 11(61.1) | 0.59 |
| Race |  |  | 0.53 |
| White (%) | 36 (34.3) | 9 (50) |  |
| Black or African American (%) | 28 (26.7) | 3 (16.7) |  |
| Asian (%) | 8 (7.6) | 2 (11.1) |  |
| Unknown/not reported (%) | 33 (31.4) | 4 (22.2) |  |
| Ethnicity |  |  | 0.09 |
| Not Hispanic or Latino (%) | 90 (85.7) | 18 (100) |  |
| Hispanic or Latino (%) | 15 (14.3) | 0 (0) |  |
| Hunt & Hess (HH) Score |  |  | 0.32 |
| 1 (%) | 1 (1.0) | 1 (5.6) |  |
| 2 (%) | 59 (56.2) | 9 (50) |  |
| 3 (%) | 23 (21.9) | 3 (16.7) |  |
| 4 (%) | 6 (5.7) | 0 (0) |  |
| 5 (%) | 16 (15.2) | 5 (27.8) |  |
| Hunt & Hess (HH) Score Dichotomized |  |  | 0.52 |
| Low-Grade (%) – 1, 2, & 3 | 83 (79.0) | 13 (72.2) |  |
| High-Grade (%) – 4 & 5 | 22 (21.0) | 5 (27.8) |  |
| Modified Fisher (mF) Score |  |  | 0.92 |
| Grade 0 (%) | 8 (7.6) | 1 (5.6) |  |
| Grade 1 (%) | 12 (11.4) | 3 (16.7) |  |
| Grade 2 (%) | 12 (11.4) | 2 (11.1) |  |
| Grade 3 (%) | 20 (19.0) | 2 (11.1) |  |
| Grade 4 (%) | 53 (50.5) | 10 (55.6) |  |
| Modified Fisher (mF) Score<br>Dichotomized |  |  | 0.69 |
| Low-Grade (%) – 0, 1, 2, & 3 | 52 (49.5) | 8 (44.4) |  |
| High-Grade (%) – 4 | 53 (50.5) | 10 (55.6) |  |
| World Federation of<br>Neurosurgical Societies<br>(WFNS) |  |  | 0.44 |
| Grade 1 (%) | 54 (51.4) | 10 (55.6) |  |
| Grade 2 (%) | 18 (17.1) | 2 (11.1) |  |
| Grade 3 (%) | 3 (2.9) | 1 (5.6) |  |
| Grade 4 (%) | 11 (10.5) | 0 (0) |  |
| Grade 5 (%) | 19 (18.1) | 5 (27.8) |  |
| EVD |  |  | 0.86 |
| No (%) | 56 (53.3) | 10 (55.6) |  |
| Yes (%) | 49 (46.7) | 8 (44.4) |  |
| Treatment Modality |  |  | 0.04 |
| No Treatment (%) | 35 (33.3) | 5 (27.8) |  |
| Clip (%) | 24 (22.9) | 0 (0) |  |
| Coil (%) | 41 (39.0) | 13 (72.2) |  |
| Other* (%) | 5 (4.8) | 0 (0) |  |
| Aneurysmal (%) | 74 (70.5) | 13 (72.2) | 0.88 |
| Average aneurysm size (mm) ± SD | 6.0 ± 3.2 | 6.5 ± 3.7 | 0.63 |
| Aneurysm Location |  |  | 0.50 |
| ICA | 3 | 1 |  |
| MCA | 11 | 4 |  |
| ACA | 4 | 0 |  |
| Ophthalmic | 3 | 1 |  |
| Vertebral | 2 | 0 |  |
| Basilar | 3 | 0 |  |
| PCA | 3 | 0 |  |
| Pcomm | 24 | 2 |  |
| PICA | 3 | 2 |  |
| Acomm | 17 | 2 |  |
| Other | 3 | 1 |  |
| Angiogram-negative | 31 | 5 |  |

\*In the other treatment category, 4 patients received onyx embolization and one received pipeline flow diversion

In the cohort of 123 patients, 10% (12/123) developed DCI, 41% (50/123) developed radiographic vasospasm, and 42% (52/123) had discharge mRS > 3 (Figure 3). Of the two patients who experienced DCI alone, both suffered CD, and of the ten patients who experienced DCI and vasospasm together, two suffered CD, one suffered CI, and seven suffered CD and CI together (Table 2). One patient who was determined to have vasospasm and DCI with both CI and CD had a baseline mRS of three due primarily to cerebral palsy. One patient who was determined to not have vasospasm or DCI had a baseline mRS of four primarily due to multiple sclerosis. There was one case of ventriculitis.

**Figure 3.**
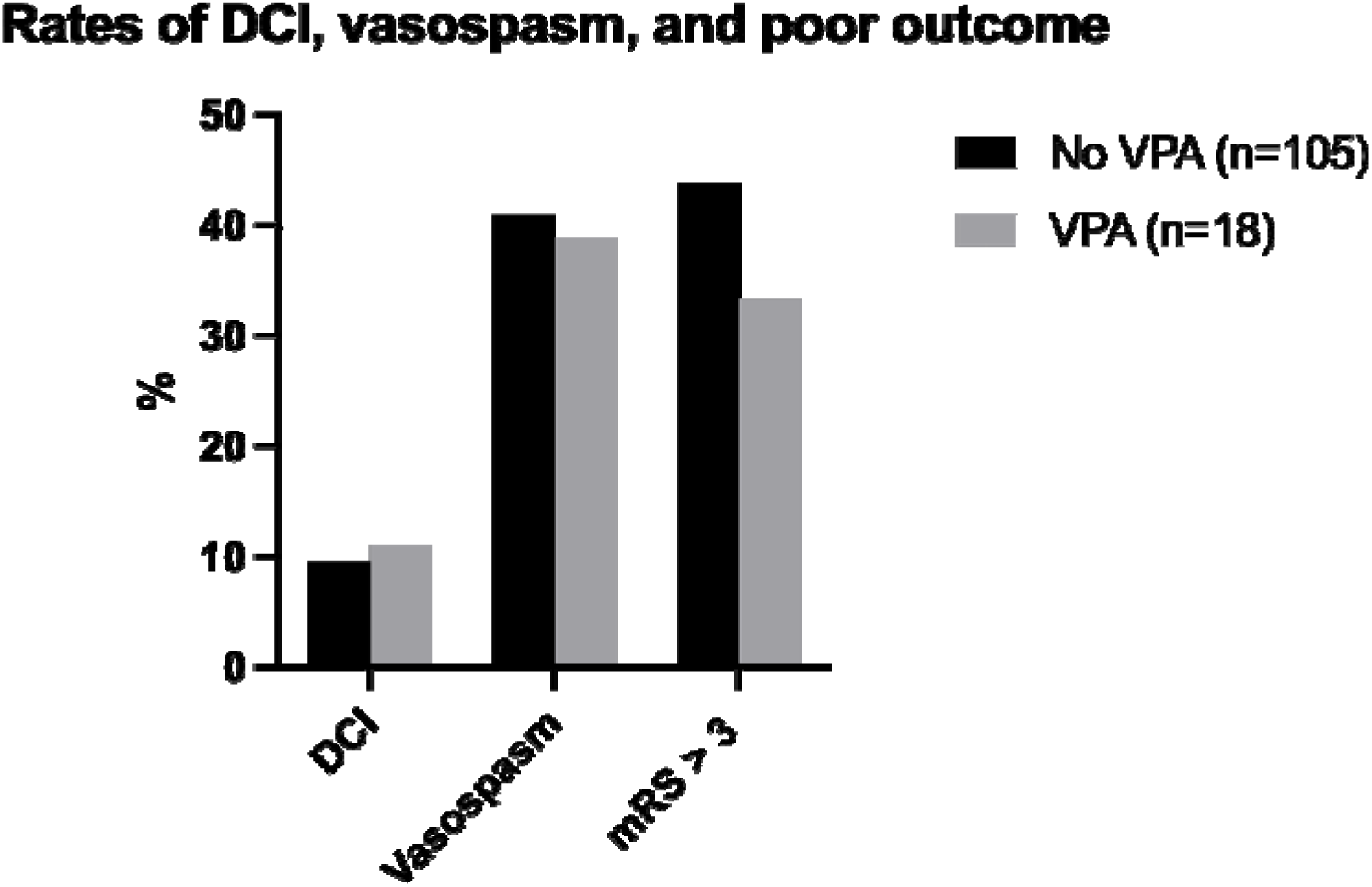
Percentages of patients who experienced DCI, vasospasm, or mRS > 3 grouped according to whether they received VPA. Percentages are shown as percentages of total patients who either did or did not receive VPA treatment.

**Table 2.**
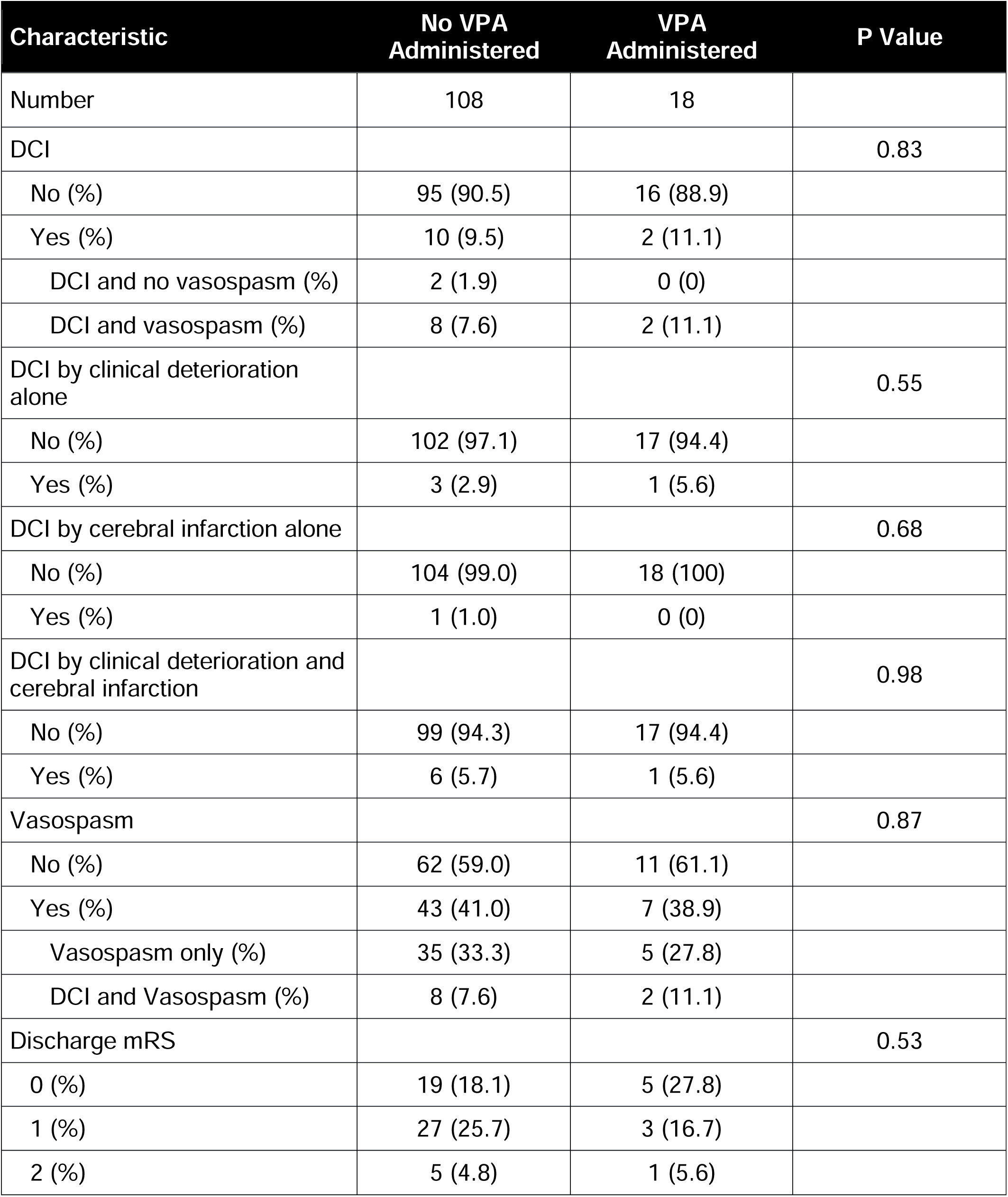
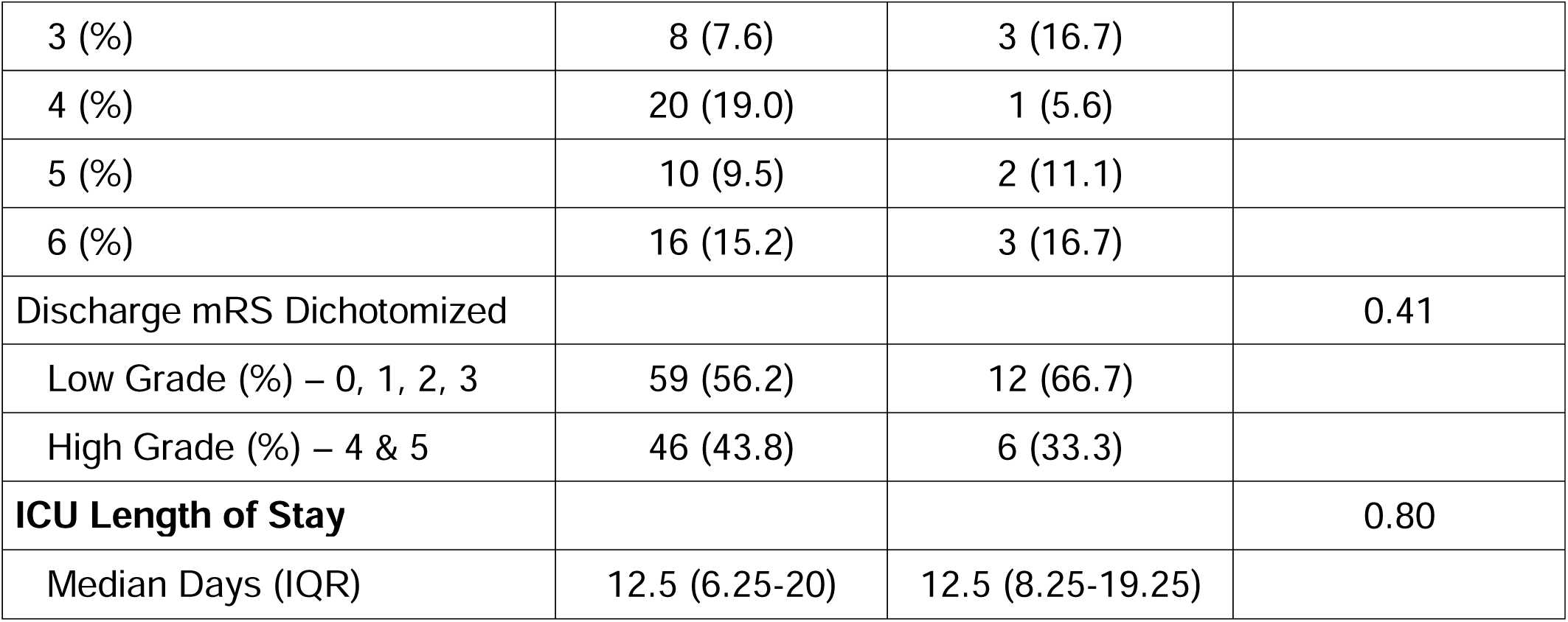
Outcomes of patients in the VPA and non-VPA groups.

Among patients who received VPA (n = 18), 11.1% had DCI and 38.9% had radiographic vasospasm, compared to 9.5% and 41.0% in the non-VPA group (n = 105). In the VPA group, none of the patients had DCI alone, 27.8% had vasospasm alone, and 11.1% had DCI and vasospasm together, compared to 1.9%, 33.3%, and 7.6% in the non-VPA group, respectively (Table 2). The median length of stay (LOS) was 12.5 days for those in both the VPA and non-VPA groups (Table 2).

Odds ratios (OR) between VPA and selected outcomes were as follows: DCI OR = 1.07 (95% CI: 0.20 – 5.80), vasospasm OR = 0.64 (95% CI: 0.19 – 1.98), and discharge mRS > 3 OR = 0.45 (95% CI: 0.10 – 1.64) after adjusting for age, HH grade > 3, and dichotomized mF (Figure 4).

**Figure 4.**
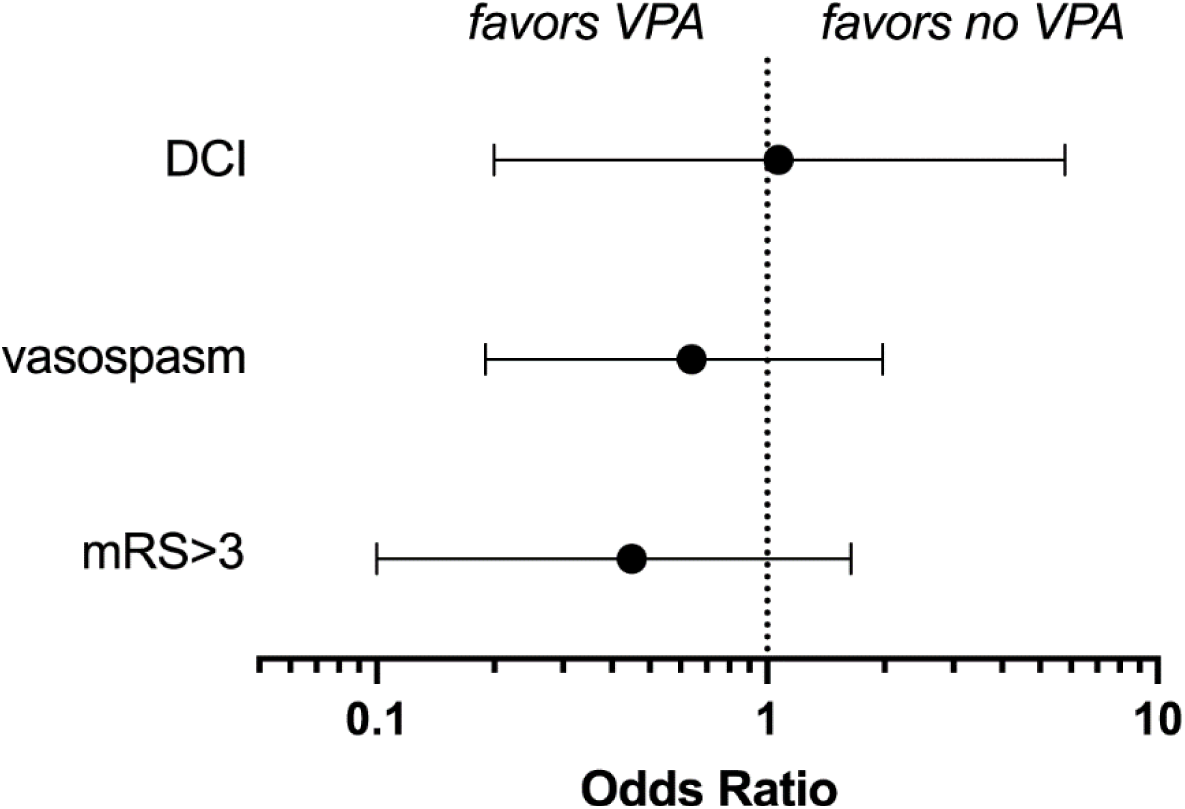
Adjusted comparison of the outcome variables in the VPA and non-VPA groups.

Younger age, (OR = 0.96, 95% CI: 0.93 - 0.99), mF score = 4 (OR = 4.14, 95% CI: 1.81 – 9.45), and HH grade > 3 (OR = 2.92, 95% CI: 1.11 – 7.69) were all associated with subsequent development of vasospasm. Older age (OR = 1.04, 95% CI: 1.01 - 1.07) and HH grade > 3 (OR = 14.5, 95% CI: 4.31 – 48.6) were associated with an increased likelihood for having a discharge mRS > 3.

We conducted a safety review of all patients who were administered VPA and did not detect VPA-related complications. Platelet levels remained stable after initiation of VPA (Figure 5). The mean ALT levels before and after VPA administration were 29.3 (SD 21) and 38.2 (SD 33.8), respectively (p = 0.12). The mean AST levels before and after VPA administration were 35.9 (SD 16.2) and 35.5 (SD 20.3), respectively (p = 0.81) (Figure 6).

**Figure 5.**
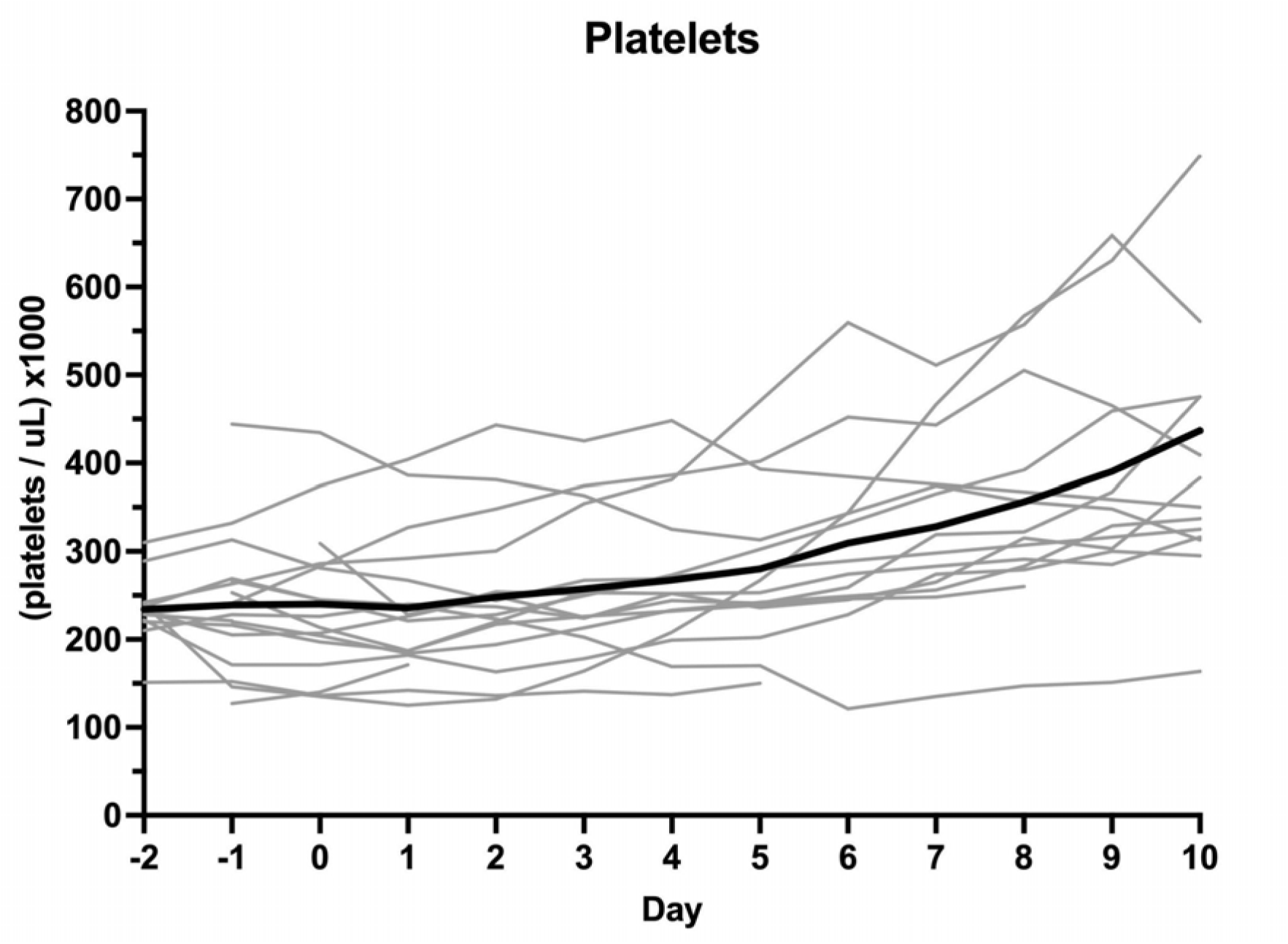
Platelet counts of all patients in the study who received VPA from the first two days before receiving VPA until ten days after VPA treatment began. Day zero measurements were the first measurements obtained after initiation of VPA treatment. The mean platelets are displayed in black and the platelet values for each individual patient are shown in gray.

**Figure 6.**
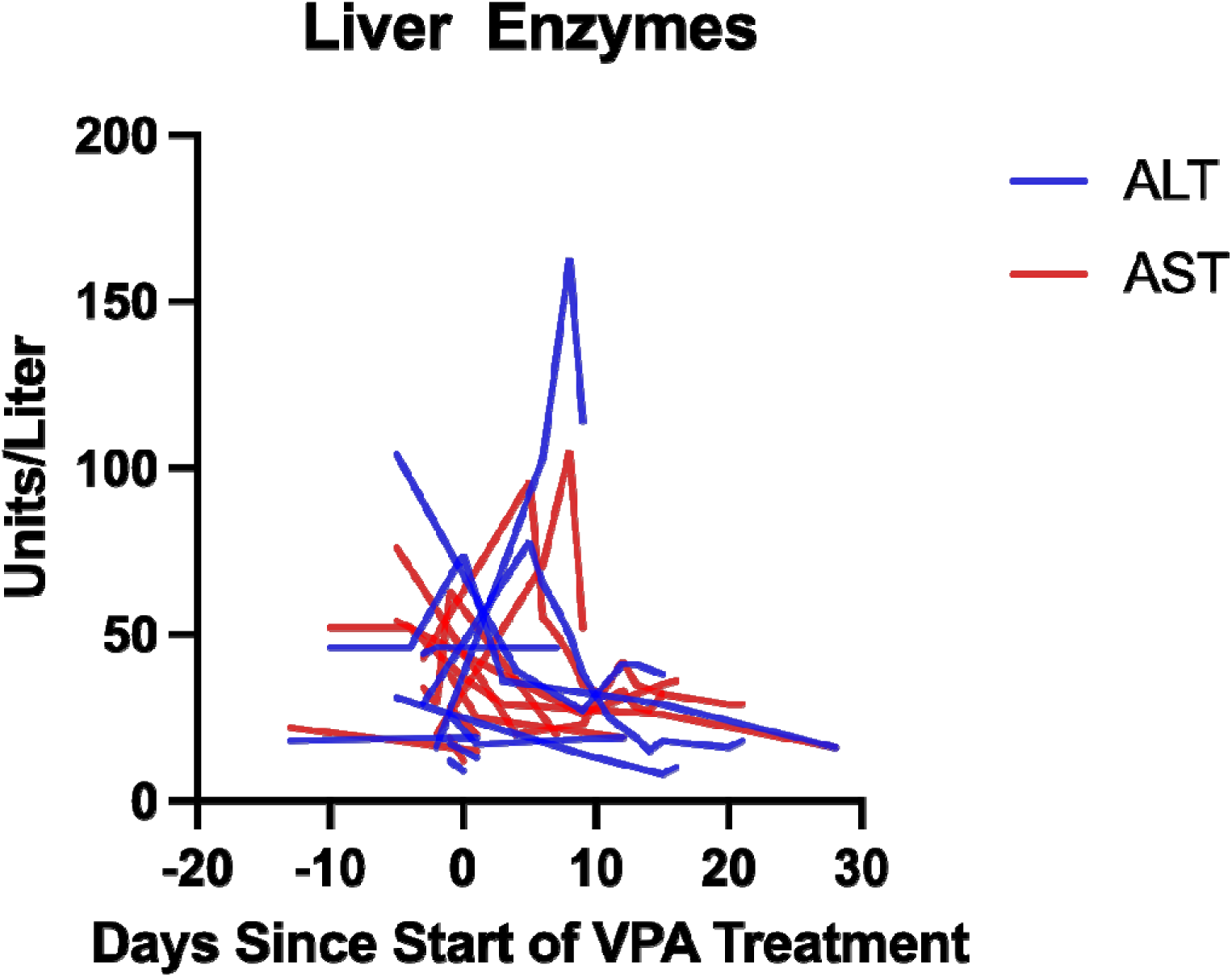
AST and ALT counts of all patients in the study who received VPA. Pre-treatment values include the closest day to the beginning of treatment and post-treatment values continue until the highest value was reached. Day zero measurements were the first measurements obtained after initiation of VPA treatment.

## Discussion

There have been no new pharmacologic therapies in the United States to improve outcomes in patients with SAH since the introduction of nimodipine in 1983.^23,24^ VPA has been shown to improve outcomes in animal models, and we sought to determine if we could detect a benefit of VPA in a human study of non-traumatic SAH. We did not see an association between VPA and the rate of DCI. However, we detected an association of VPA with reduced radiographic vasospasm and better discharge mRS which did not reach the threshold for statistical significance. Importantly, VPA use in endovascularly coiled patients appears safe.

Animal studies have shown that valproic acid (VPA) improves outcome after SAH through a mechanism of aborting CSD-mediated secondary injury.^12,13,25^ To our knowledge, this is the first study evaluating the effect of VPA on SAH outcomes in humans. Reasons for not seeing an effect of VPA in our patient population includes the absence of the invasive intracranial monitoring necessary to determine which patients developed CSDs. Future studies could benefit from measurement of CSDs in patients. Currently, invasive subdural electrocorticography (ECoG) is required to reliably measure CSDs in humans; however, there is emerging work to detect CSD correlates with electroencephalography (EEG).^26,27^ VPA might indeed suppress CSDs in humans like it does in rodents, but until we are able to reliably, safely, and directly measure CSDs in humans, it will be difficult to demonstrate this effect. Future studies should aim to find better ways of non-invasive measurement of CSDs in humans once such approaches become more widespread.

VPA may exert neuroprotective effects through mechanisms other than CSDs. Previous studies involving SAH rat models have shown that VPA reduced DCI-induced neuro-apoptosis by upregulating histone H3 acetylation and Akt phosphorylation^28^ and decreased blood brain barrier disruption and cerebral edema by modulating HSP70/MMPs and HSP70/Akt pathways.^29^ In pig models of TBI and hemorrhagic shock, VPA decreased lesion size, reduced neurologic impairment,^30^ decreased neuro-apoptosis, inflammation, and degenerative changes.^31^ Therefore, there are potential CSD-mediation and CSD-independent pathways through which VPA could exert its effects.

The limitations of our paper include those common to other retrospective studies including selection bias. Patients were from a single center. It is notable that none of the patients who received VPA had undergone surgical clipping procedures for aneurysm treatment. VPA has historically been avoided in SAH due to concerns about bleeding complications in the setting of a craniotomy. In the era of endovascular treatment of ruptured aneurysms VPA is now being used more frequently in patients with SAH for agitation, headache, and seizure prophylaxis. At the same time, the use of VPA in some patients may reflect a variation in clinical management of SAH patients such that VPA might have been administered for different reasons based on clinician preference and outcomes might vary based on differing clinical management.

We demonstrate that VPA is safe for use in patients with aneurysmal subarachnoid hemorrhage treated with endovascular coiling. We did not detect statistically significant effects on development of DCI, radiographic vasospasm, and discharge mRS > 3 which may be due to low sample size. A future adequately powered multicenter study to determine the effect of VPA on DCI and long-term outcomes in nontraumatic SAH patients is justified.

## Abbreviations

SAH: subarachnoid hemorrhage
DCI: delayed cerebral ischemia
DSA: digital subtraction angiography
CTA: computed tomography angiography
TCD: transcranial doppler sonography
SD: spreading depolarization
GCS: Glasgow Coma Scale
VPA: valproic acid
AVM: arteriovenous malformation
RCVS: reversible cerebral vasoconstriction syndrome
BMC: Boston Medical Center
LOS: length of stay
ECoG: electrocorticography
EEG: electroencephalography
CMO: comfort measures only

## Acknowledgements

This work was funded by the National Institutes of Health (K08NS112601 and R01NS136224 to DYC; K23NS116033 to CO, R01NS102574 and R01NS128342 to DG), the Andrew David Heitman Foundation (DYC), a Boston University Undergraduate Research Opportunity Program award (JY), and the Grinspoon Family Award (AD). This publication was also supported by the National Center for Advancing Translational Sciences, National Institutes of Health, through BU-CTSI Grant Number 1UL1TR001430. Its contents are solely the responsibility of the authors and do not necessarily represent the official views of the NIH. We thank Cenk Ayata for helpful comments on the manuscript.

## Data availability

The data of this study are available from the corresponding author upon request.

## References

1. Muehlschlegel S. Subarachnoid Hemorrhage. Continuum (Minneap Minn). 2018;24:1623–1657. doi: 10.1212/CON.0000000000000679

2. Connolly ES, Jr., Rabinstein AA, Carhuapoma JR, Derdeyn CP, Dion J, Higashida RT, Hoh BL, Kirkness CJ, Naidech AM, Ogilvy CS, et al. Guidelines for the management of aneurysmal subarachnoid hemorrhage: a guideline for healthcare professionals from the American Heart Association/American Stroke Association. Stroke; a journal of cerebral circulation. 2012;43:1711–1737. doi: 10.1161/STR.0b013e3182587839

3. SVIN COVID-19 Global SAH Registry. Global impact of the COVID-19 pandemic on subarachnoid haemorrhage hospitalisations, aneurysm treatment and in-hospital mortality: 1-year follow-up. J Neurol Neurosurg Psychiatry. 2022. doi: 10.1136/jnnp-2022-329200

4. Frontera JA, Fernandez A, Schmidt JM, Claassen J, Wartenberg KE, Badjatia N, Connolly ES, Mayer SA. Defining vasospasm after subarachnoid hemorrhage: what is the most clinically relevant definition? Stroke; a journal of cerebral circulation. 2009;40:1963–1968. doi: 10.1161/STROKEAHA.108.544700

5. Vergouwen MD, Vermeulen M, van Gijn J, Rinkel GJ, Wijdicks EF, Muizelaar JP, Mendelow AD, Juvela S, Yonas H, Terbrugge KG, et al. Definition of delayed cerebral ischemia after aneurysmal subarachnoid hemorrhage as an outcome event in clinical trials and observational studies: proposal of a multidisciplinary research group. Stroke; a journal of cerebral circulation. 2010;41:2391–2395. doi: 10.1161/STROKEAHA.110.589275

6. Chung DY, Abdalkader M, Nguyen TN. Aneurysmal Subarachnoid Hemorrhage. Neurol Clin. 2021;39:419–442. doi: 10.1016/j.ncl.2021.02.006

7. Chung DY, Oka F, Ayata C. Spreading Depolarizations: A Therapeutic Target Against Delayed Cerebral Ischemia After Subarachnoid Hemorrhage. Journal of clinical neurophysiology : official publication of the American Electroencephalographic Society. 2016;33:196–202. doi: 10.1097/WNP.0000000000000275

8. Sugimoto K, Chung DY. Spreading Depolarizations and Subarachnoid Hemorrhage. Neurotherapeutics. 2020;17:497–510. doi: 10.1007/s13311-020-00850-5

9. Dreier JP, Petzold G, Tille K, Lindauer U, Arnold G, Heinemann U, Einhaupl KM, Dirnagl U. Ischaemia triggered by spreading neuronal activation is inhibited by vasodilators in rats. J Physiol. 2001;531:515–526. doi: 10.1111/j.1469-7793.2001.0515i.x

10. Dreier JP, Windmuller O, Petzold G, Lindauer U, Einhaupl KM, Dirnagl U. Ischemia triggered by red blood cell products in the subarachnoid space is inhibited by nimodipine administration or moderate volume expansion/hemodilution in rats. Neurosurgery. 2002;51:1457–1465; discussion 1465-1457.

11. Sugimoto K, Nomura S, Shirao S, Inoue T, Ishihara H, Kawano R, Kawano A, Oka F, Suehiro E, Sadahiro H, et al. Cilostazol decreases duration of spreading depolarization and spreading ischemia after aneurysmal subarachnoid hemorrhage. Annals of neurology. 2018;84:873–885. doi: 10.1002/ana.25361

12. Hamming AM, van der Toorn A, Rudrapatna US, Ma L, van Os HJ, Ferrari MD, van den Maagdenberg AM, van Zwet E, Poinsatte K, Stowe AM, et al. Valproate Reduces Delayed Brain Injury in a Rat Model of Subarachnoid Hemorrhage. Stroke; a journal of cerebral circulation. 2017;48:452–458. doi: 10.1161/STROKEAHA.116.014738

13. Tso MK, Lass E, Ai J, Loch Macdonald R. Valproic Acid treatment after experimental subarachnoid hemorrhage. Acta neurochirurgica Supplement. 2015;120:81–85. doi: 10.1007/978-3-319-04981-6_14

14. von Elm E, Altman DG, Egger M, Pocock SJ, Gøtzsche PC, Vandenbroucke JP. The Strengthening the Reporting of Observational Studies in Epidemiology (STROBE) statement: guidelines for reporting observational studies. J Clin Epidemiol. 2008;61:344–349. doi: 10.1016/j.jclinepi.2007.11.008

15. Zafar SF, Westover MB, Gaspard N, Gilmore EJ, Foreman BP, O’Connor KL, Rosenthal ES. Interrater Agreement for Consensus Definitions of Delayed Ischemic Events After Aneurysmal Subarachnoid Hemorrhage. Journal of clinical neurophysiology : official publication of the American Electroencephalographic Society. 2016;33:235–240. doi: 10.1097/WNP.0000000000000276

16. Flanagin A, Frey T, Christiansen SL, Bauchner H. The Reporting of Race and Ethnicity in Medical and Science Journals: Comments Invited. JAMA. 2021;325:1049–1052. doi: 10.1001/jama.2021.2104

17. Gupta S, Fenves AZ, Hootkins R. The Role of RRT in Hyperammonemic Patients. Clin J Am Soc Nephrol. 2016;11:1872–1878. doi: 10.2215/cjn.01320216

18. Anderson GD, Lin YX, Berge C, Ojemann GA. Absence of bleeding complications in patients undergoing cortical surgery while receiving valproate treatment. J Neurosurg. 1997;87:252–256. doi: 10.3171/jns.1997.87.2.0252

19. Kurwale N, Garg K, Arora A, Chandra PS, Tripathi M. Valproic acid as an antiepileptic drug: Is there a clinical relevance for the epilepsy surgeon? Epilepsy Res. 2016;127:191–194. doi: 10.1016/j.eplepsyres.2016.09.005

20. Williamson DR, Albert M, Heels-Ansdell D, Arnold DM, Lauzier F, Zarychanski R, Crowther M, Warkentin TE, Dodek P, Cade J, et al. Thrombocytopenia in critically ill patients receiving thromboprophylaxis: frequency, risk factors, and outcomes. Chest. 2013;144:1207–1215. doi: 10.1378/chest.13-0121

21. van der Steen WE, Leemans EL, van den Berg R, Roos Y, Marquering HA, Verbaan D, Majoie C. Radiological scales predicting delayed cerebral ischemia in subarachnoid hemorrhage: systematic review and meta-analysis. Neuroradiology. 2019;61:247–256. doi: 10.1007/s00234-019-02161-9

22. Yu H, Zhan R, Wen L, Shen J, Fan Z. The relationship between risk factors and prognostic factors in patients with shunt-dependent hydrocephalus after aneurysmal subarachnoid hemorrhage. J Craniofac Surg. 2014;25:902–906. doi: 10.1097/scs.0000000000000561

23. Allen GS, Ahn HS, Preziosi TJ, Battye R, Boone SC, Boone SC, Chou SN, Kelly DL, Weir BK, Crabbe RA, et al. Cerebral arterial spasm--a controlled trial of nimodipine in patients with subarachnoid hemorrhage. The New England journal of medicine. 1983;308:619–624. doi: 10.1056/NEJM198303173081103

24. Pickard JD, Murray GD, Illingworth R, Shaw MD, Teasdale GM, Foy PM, Humphrey PR, Lang DA, Nelson R, Richards P, et al. Effect of oral nimodipine on cerebral infarction and outcome after subarachnoid haemorrhage: British aneurysm nimodipine trial. Bmj. 1989;298:636–642.

25. Ayata C, Jin H, Kudo C, Dalkara T, Moskowitz MA. Suppression of cortical spreading depression in migraine prophylaxis. Annals of neurology. 2006;59:652–661. doi: 10.1002/ana.20778

26. Drenckhahn C, Winkler MK, Major S, Scheel M, Kang EJ, Pinczolits A, Grozea C, Hartings JA, Woitzik J, Dreier JP. Correlates of spreading depolarization in human scalp electroencephalography. Brain. 2012;135:853–868. doi: 10.1093/brain/aws010

27. Chamanzar A, Elmer J, Shutter L, Hartings J, Grover P. Noninvasive and reliable automated detection of spreading depolarization in severe traumatic brain injury using scalp EEG. Commun Med (Lond). 2023;3:113. doi: 10.1038/s43856-023-00344-3

28. Wu CH, Tsai YC, Tsai TH, Kuo KL, Su YF, Chang CH, Lin CL. Valproic Acid Reduces Vasospasm through Modulation of Akt Phosphorylation and Attenuates Neuronal Apoptosis in Subarachnoid Hemorrhage Rats. International journal of molecular sciences. 2021;22. doi: 10.3390/ijms22115975

29. Ying GY, Jing CH, Li JR, Wu C, Yan F, Chen JY, Wang L, Dixon BJ, Chen G. Neuroprotective Effects of Valproic Acid on Blood-Brain Barrier Disruption and Apoptosis-Related Early Brain Injury in Rats Subjected to Subarachnoid Hemorrhage Are Modulated by Heat Shock Protein 70/Matrix Metalloproteinases and Heat Shock Protein 70/AKT Pathways. Neurosurgery. 2016;79:286–295. doi: 10.1227/NEU.0000000000001264

30. Wakam GK, Biesterveld BE, Pai MP, Kemp MT, O’Connell RL, Williams AM, Srinivasan A, Chtraklin K, Siddiqui AZ, Bhatti UF, et al. Administration of valproic acid in clinically approved dose improves neurologic recovery and decreases brain lesion size in swine subjected to hemorrhagic shock and traumatic brain injury. J Trauma Acute Care Surg. 2021;90:346–352. doi: 10.1097/TA.0000000000003036

31. Chang P, Williams AM, Bhatti UF, Biesterveld BE, Liu B, Nikolian VC, Dennahy IS, Lee J, Li Y, Alam HB. Valproic Acid and Neural Apoptosis, Inflammation, and Degeneration 30 Days after Traumatic Brain Injury, Hemorrhagic Shock, and Polytrauma in a Swine Model. J Am Coll Surg. 2019;228:265–275. doi: 10.1016/j.jamcollsurg.2018.12.026

